# Autoimmune side-effect of immunotherapy in lung cancer treatment revealed from large-scale cohort

**DOI:** 10.1101/2024.12.03.24318450

**Authors:** Yan Sun, Shihao Yang

**Affiliations:** H. Milton Stewart School of Industrial and Systems Engineering, Georgia Institute of Technology, 755 Ferst Dr NW, Atlanta, 30318, Georgia, USA

**Keywords:** lung cancer, immunology, causal inference, survival analysis, autoimmune, chemotherapy

## Abstract

Although immune checkpoint inhibitors have illustrated strong benefits in patient survival and have been widely acknowledged in treating lung cancer, they may be subject to increased risk of immune-related adverse effects (irAEs). Although existing literature have studied the mechanisms of irAEs of immunotherapy, it is difficult to quantify such effect, especially at a large-scale real-world population level. In this paper, the autoimmune-related risk of multiple immune checkpoint inhibitors is compared with that of chemotherapy based on Medicaid and CHIP TAF (T-MSIS Analytic File) data of over 100,000 patient samples from 2012 to 2018. Results show that the irAEs of immunotherapy is significantly higher than chemotherapy in both unadjusted and adjusted samples from the dataset. Analysis on subpopulation and specific disease types further shows that certain immunotherapy treatments are associated with higher risk of irAEs, and the risk of certain autoimmune diseases may vary. We also illustrate the robustness of our conclusion through additional sensitivity analysis, confirming the necessity of keeping track of autoimmune side effects of immune checkpoint inhibitors for medicine researchers. Our methods are also available to evaluate effectiveness and side effects of novel therapies at a large-scale population level.

## 1 Introduction

Immune checkpoint inhibitors have received phenomenal clinical success in treating multiple types of cancer, and have innovated the landscape of modern cancer therapy [1]. Since the approval of immune checkpoint inhibitor, Ipilimumab, by the U.S. Food and Drug Administration (FDA), many types of immunotherapy have been discovered for specific cancer types. The most common immunotherapies in recent years for cancer treatment are immune checkpoints T-lymphocyte-associated protein 4 (CTLA-4) inhibitors and programmed cell death protein 1 (PD-1) inhibitors [2]. There are also other types of immune checkpoint inhibitors, such as LAG-3 inhibitors.

Unlike conventional mechanism of chemotherapy, immunotherapy enables certain aspects of immune system to eliminate targeted tumor cells [3]. CTLA-4 inhibitor binds to CTLA-4 receptor, blocks the expression of CTLA-4 protein, thus preventing its attachment to B7 receptor in APCs and decreasing the activation threshold of T cells, increasing immune responses against tumor cells [4]. Programmed cell death protein 1 (PD-1) inhibitors, including Pembrolizumab (Keytruda ™) and Nivolumab (Opdivo ™), reactivate the host immune response against cancer through suppression of PD-1, which is a transmembrane protein mediating intracellular signaling events during T cell activation [5], and is crucial for the maintenance of peripheral tolerance and containing immune responses to avoid immunopathology [6]. The ligands of PD-1, PD-L1 and PD-L2, both found on the surface of antigen-presenting cells, also lead to the development of novel immunotherapy, such as Atezolizumab and Avelumab. Both PD-1 and PD-L1 inhibitors have been proven helpful in treating many different types of cancer [7]. In addition to the substantial treatment effects and survival gains of immune checkpoint inhibitors [8, 9], immunotherapy avoids common cytotoxic damage to non-tumor cells due to conventional radiotherapy and chemotherapy [10], and has gained increasing clinical popularity for various types of cancer, such as lung cancer, breast cancer [11] and melanoma [2]. Currently several common immunotherapies have been applied in lung cancer treatment: Nivolumab (Opdivo ™), Pembrolizumab (Keytruda ™), Ipilimumab (Yervoy ™) and Atezolizumab (Tecentriq ™). Novel immunotherapy drugs are also being invented in recent years, such as Dostarlimab, but are either not formally approved or less widely applied due to recency.

Although the functionality has been proven effective [6], immune checkpoint inhibitors may also lead to certain side effects [12]. Common adverse effects of immunotherapy include diarrhea, fatigue, nausea, decreased appetite [13] and neurologic complications [10]. Besides, by targeting a checkpoint protein, the inhibitors may remove certain safeguards on the immune system. Correspondingly, immune system may attack other parts of the body, which can cause serious reactions, known as autoimmune-related adverse effects (irAEs) [14]. Common irAEs including hypothyroidism and thyroditis have been reported in clinical application of immunotherapy clinical trials and small-size observational studies [15]. However, irAEs are still not well explored at a large-population level due to the recency of clinical application [16]. Moreover, although most patients under immunotherapy are reported with irAEs, some adverse effects may have much lower incidence rate, which makes large-scale observational studies necessary.

Recent advancement of electronic health data (EHR) has facilitated studies on evaluating adverse events of novel treatment strategies based on large-population scale real-world data (RWD) [17]. RWD is a source of information related to patient health conditions and healthcare delivery [18], often collected routinely by hospitals or healthcare agencies. Common sources of RWD include nationwide insurance datasets, hospital medical records and medicaid data. One advantage of RWD is accumulation of sufficient evidence of side effects with smaller probabilities to dissect issues pertaining to multiple sectors [19]. Moreover, observational data sources such as health claims [20] are usually population based, longitudinally collected and structurally constructed, which facilitates large-scale studies.

However, real world data may also face several challenges. One major drawback of using RWD is the difficulty in justifying causality. Different from experimental data, RWD is collected without specific research purpose, thus identification the treatment effects may be challenging. RWD may also incorporate incomplete or missing data, stimulated reporting and duplicate reporting. To address such issues, we employed matching method from the causal inference literature to reduce the biased relationship between immunotherapy and subsequent irAEs, therefore establishing a strengthened causal effect.

The scope of this paper focuses on evaluation of autoimmune risk of immune checkpoint therapy in lung cancer treatment. Lung cancer has been the primary cause of cancer-related mortality leading to over 1.5 million deaths per year worldwide [21], and is widely known to be challenging for immunotherapy treatments [22]. Despite the success of several immunotherapies in certain cancers, immunotherapy have historically failed in lung cancer [22]. However, recent studies have demonstrated significant survival benefit of immunotherapy in treating lung cancer [23]. This paper focuses on a comprehensive cohort study of the immune-related adverse effect of immunotherapy on lung cancer patients, utilizing the real world dataset of United States Medicaid data, in comparison to irAEs of traditional chemotherapy.

The paper is organized as follows: in Sec 2.2 we present the workflow of data processing and visualize the number of patients (records) after each procedure, based on inclusion-exclusion criteria (see Sec 2.5). The matching methodology is introduced in Sec 3, and the main results are provided in Sec 4, where Kaplan Meier curves are provided and log-rank test for the hazard ratio is conducted along with weighted cumulative incidence ratio. Sec 4.3 shows the results of sensitivity analysis, where we investigate whether the risk increase for specific irAEs and immunotherapy (Nivolumab, Pembrolizumab, Ipilimumab and Atezolizumab). Additional analysis using different sub-population and matching criteria are provided in Sec 4.4. We wrap up the study with discussions in Sec 5.

## 2 Materials and methods

### 2.1 Overview of the CMS dataset

Our study utilizes the RWD of de-identified medicaid claim data from The Centers for Medicare and Medicaid services (CMS). Medicaid is a joint federal and state program that helps cover medical costs for patients with limited income and resources. The medicaid dataset contains the medicaid claims from patients prior to December 31, 2018, with a total of more than 500,000 unique patients with 30+ million records. **Written informed consent have been acquired from the patients before the enrollment of medical plan for research purposes, and the consent forms are required for each patient to receive chemo- or immunotherapy treatment**. The claims records provide diagnosis codes encoded by the International Statistical Classification of Diseases and Related Health Problems, in mixed ninth (ICD-9) and tenth edition (ICD-10). Treatment procedures are encoded according to Current Procedural Terminology and Healthcare Common Procedure Coding System (HCPCS). Details of the treatments such as date and dosage are also provided. In particular, the diagnosis date of lung cancer and earliest irAEs are listed, which enables investigation of the causal relationship between immune inhibitors and irAEs.

### 2.2 Study cohort

The summary of dataset after each data processing procedure is visualized in Fig.1 to show how much data is selected, merged and filtered in each step. 5725 qualified patients with immunotherapy and 102490 qualified patients with chemotherapy are finally acquired as the targeted population of the study. Table 1 provides the detailed demographics after the data filtering procedure.

**Fig. 1:**
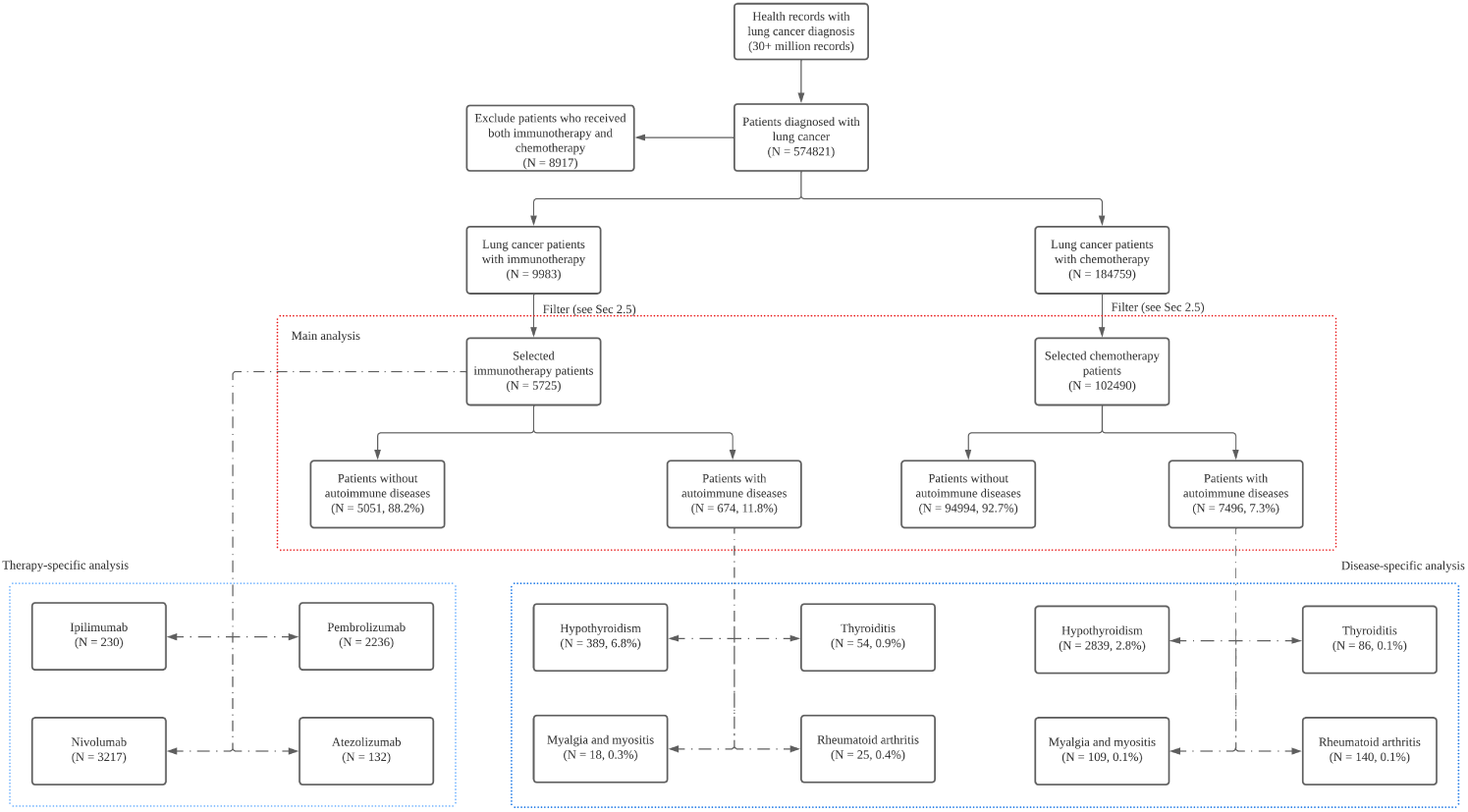
Diagram of filtering the patient samples.A patient may receive combined immunotherapy and exhibit multiple syndromes after treatment.

**Table 1:** Patient characteristics.

|  |  | Ipilimumab |  | Nivolumab |  | Pembrolizumab |  | Atezolizumab |  | Immunotherapy |  | Chemotherapy |  |
| --- | --- | --- | --- | --- | --- | --- | --- | --- | --- | --- | --- | --- | --- |
|  |  | N<sup>1</sup> | p<sup>2</sup> | N | p | N | p | N | p | N | p | N | p |
| Total |  | 230 | 4.0% | 3217 | 56.2% | 2236 | 39.1% | 132 | 2.3% | 5725<sup>3</sup> | - | 102490 | - |
| Gender | Female | 100 | 43.5% | 1501 | 46.7% | 1102 | 49.3% | 55 | 41.7% | 2706 | 47.3% | 48495 | 47.3% |
|  | Male | 125 | 54.3% | 1653 | 51.4% | 1111 | 49.7% | 76 | 57.6% | 2926 | 51.1% | 50731 | 49.5% |
|  | Unknown | 5 | 2.1% | 63 | 2.0% | 23 | 1.0% | 1 | 0.8% | 93 | 1.6% | 3264 | 3.2% |
| Race and ethnicity | White | 132 | 57.4% | 1539 | 47.8% | 1098 | 49.1% | 62 | 47.0% | 2781 | 48.6% | 54160 | 52.9% |
|  | Black | 15 | 6.5% | 473 | 14.7% | 366 | 16.4% | 24 | 18.2% | 876 | 15.3% | 18581 | 18.1% |
|  | American Indian | 5 | 2.2% | 98 | 3.0% | 124 | 5.5% | 10 | 7.6% | 231 | 4.0% | 1293 | 1.3% |
|  | Asian | 2 | 0.9% | 28 | 8.7% | 14 | 0.6% | 0 | 0 | 44 | 0.8% | 1794 | 1.8% |
|  | Hispanic | 1 | 0.4% | 9 | 0.3% | 18 | 0.8% | 0 | 0 | 30 | 0.5% | 3065 | 3.0% |
|  | Native Hawaiian | 1 | 0.4% | 4 | 0.1% | 3 | 0.1% | 0 | 0 | 8 | 0.1% | 438 | 0.4% |
|  | Two or more races | 17 | 7.4% | 221 | 6.9% | 150 | 6.7% | 9 | 6.8% | 383 | 6.7% | 2923 | 2.9% |
|  | Unknown | 57 | 24.8% | 845 | 26.3% | 463 | 20.7% | 27 | 20.5% | 1372 | 24.0% | 20236 | 19.7% |
| Age | [0, 30) | 10 | 4.3% | 15 | 0.5% | 8 | 0.4% | 1 | 0.8% | 31 | 0.5% | 928 | 0.9% |
|  | [30, 40) | 12 | 5.2% | 39 | 1.2% | 17 | 0.8% | 3 | 2.3% | 65 | 1.1% | 2291 | 2.2% |
|  | [40, 50) | 47 | 20.4% | 273 | 8.5% | 125 | 5.6% | 9 | 6.8% | 434 | 7.6% | 15200 | 14.8% |
|  | [50, 60) | 87 | 37.8% | 1130 | 35.1% | 666 | 29.8% | 41 | 31.1% | 1884 | 32.9% | 42147 | 41.1% |
|  | [60, 70) | 55 | 23.9% | 1018 | 31.6% | 717 | 32.1% | 39 | 29.5% | 1813 | 31.7% | 28498 | 27.8% |
|  | [70, 80) | 14 | 6.1% | 508 | 15.8% | 444 | 19.9% | 20 | 15.2% | 980 | 17.1% | 9909 | 9.7% |
|  | [80, 100) | 4 | 1.7% | 196 | 6.1% | 256 | 11.4% | 19 | 14.4% | 476 | 8.3% | 2196 | 2.1% |
|  | Unknown | 1 | 0.4% | 38 | 1.2% | 3 | 0.1% | 0 | 0 | 42 | 0.7% | 1321 | 1.3% |
| Post-matching size |  | 226 | 97.4% | 3165 | 98.4% | 2207 | 98.7% | 128 | 97.0% | 5681 | 98.5% |  |  |

**Table 2:**
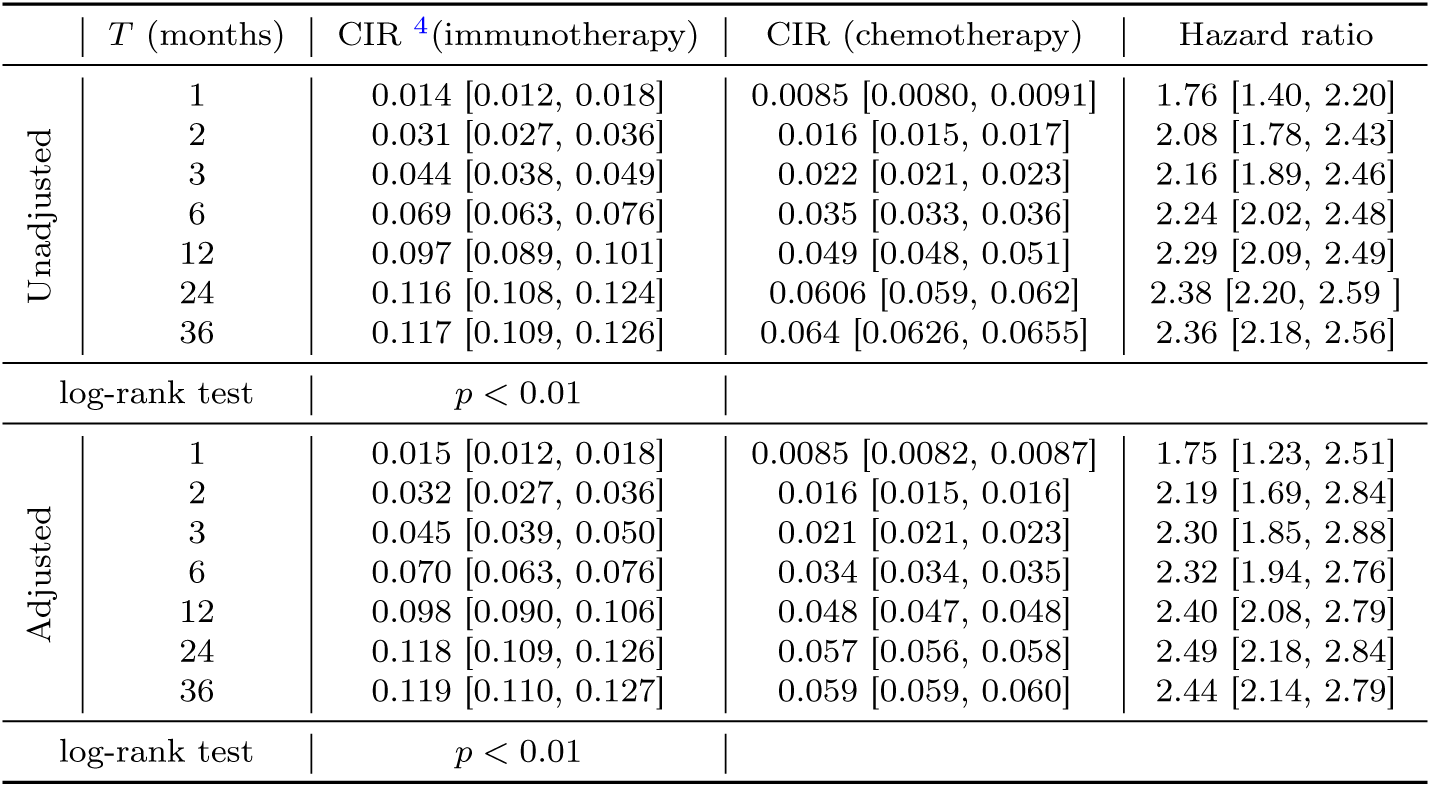
Cumulative incidence rate and hazard ratio.

| | $T$ (months) | CIR <sup>4</sup>(immunotherapy) | CIR (chemotherapy) | Hazard ratio |
| --- | --- | --- | --- | --- |
| Unadjusted | 1 | 0.014 [0.012, 0.018] | 0.0085 [0.0080, 0.0091] | 1.76 [1.40, 2.20] |
|  | 2 | 0.031 [0.027, 0.036] | 0.016 [0.015, 0.017] | 2.08 [1.78, 2.43] |
|  | 3 | 0.044 [0.038, 0.049] | 0.022 [0.021, 0.023] | 2.16 [1.89, 2.46] |
|  | 6 | 0.069 [0.063, 0.076] | 0.035 [0.033, 0.036] | 2.24 [2.02, 2.48] |
|  | 12 | 0.097 [0.089, 0.101] | 0.049 [0.048, 0.051] | 2.29 [2.09, 2.49] |
|  | 24 | 0.116 [0.108, 0.124] | 0.0606 [0.059, 0.062] | 2.38 [2.20, 2.59] |
|  | 36 | 0.117 [0.109, 0.126] | 0.064 [0.0626, 0.0655] | 2.36 [2.18, 2.56] |
| log-rank test | | $p < 0.01$ | | |
| Adjusted | 1 | 0.015 [0.012, 0.018] | 0.0085 [0.0082, 0.0087] | 1.75 [1.23, 2.51] |
|  | 2 | 0.032 [0.027, 0.036] | 0.016 [0.015, 0.016] | 2.19 [1.69, 2.84] |
|  | 3 | 0.045 [0.039, 0.050] | 0.021 [0.021, 0.023] | 2.30 [1.85, 2.88] |
|  | 6 | 0.070 [0.063, 0.076] | 0.034 [0.034, 0.035] | 2.32 [1.94, 2.76] |
|  | 12 | 0.098 [0.090, 0.106] | 0.048 [0.047, 0.048] | 2.40 [2.08, 2.79] |
|  | 24 | 0.118 [0.109, 0.126] | 0.057 [0.056, 0.058] | 2.49 [2.18, 2.84] |
|  | 36 | 0.119 [0.110, 0.127] | 0.059 [0.059, 0.060] | 2.44 [2.14, 2.79] |
| log-rank test | | $p < 0.01$ | | |

### 2.3 Immunotherapy and chemotherapy identification

We extracted the lists of immunotherapy and chemotherapy HCPCS codes for lung cancer, while other types of treatment for cancer are omitted, such as radio therapy and targeted therapy. The complete list of common CPT/HCPCS codes for immunotherapy is shown in Table 3, and chemotherapy codes are provided in the Supplementary Material Table S1. The most common immunotherapy treatments in the dataset are Nivolumab and Pembrolizumab, while Atezolizumab and Ipilimumab are also common therapies for lung cancer. Other immunotherapy treatments have very limited sample size in the dataset due to recency, and are categorized as other therapies.

**Table 3:** ICD code list of immunotherapies incorporated in the study.

| Immunotherapy | Treatment mechanism | HCPCS Code |
| --- | --- | --- |
| Ipilimumab | CTLA-4 Inhibitor | J9228 |
| Nivolumab | PD-1 Inhibitor | J9299 |
| Pembrolizumab | PD-1 Inhibitor | J9271 |
| Atezolizumab | PD-L1 Inhibitor | J9022 |
| Durvalumab | PD-L1 Inhibitor | J9173 |
| Avelumab | PD-L1 Inhibitor | J9023 |

### 2.4 Inclusion criteria

In this study, patients with at least two lung-cancer diagnosis records with chemotherapy or immunotherapy records are included, and their entire treatment histories are extracted from the database. The lung cancer mainly includes small-cell, non-small cell cancer and carcinoid, all of which can be identified through PheWAS codes and descriptions. Patients with any ICD records from Table S4 after lung cancer treatment are considered as having an irAE event. with the time-to-event length defined as the time difference from therapy initiation date to the date of first irAE diagnosis.

### 2.5 Exclusion criteria

To decrease the possibility of coding mistakes and rule out other factors which may induce biased conclusion, multiple exclusion rules were applied to filter out disqualified patients.

1. To rule out patients with potential treatment from other sources, patients whose first record is less than 30 days before the first lung cancer therapy shall be excluded from the study, following existing literature [24]. This filtering procedure is to eliminate patients with insufficient treatment history in the database, which may indicate prior cancer treatment or irAE events. In the main analysis, the quiescence time is set as 30 days, and in the sensitivity analysis different quiescence time will be discussed.
2. The first therapy for cancer the patients shall be prior to the first date of diagnosis with immune related adverse events, as there is little evidence that shows the causal relationship between therapy and autoimmune effects.
3. The patients that receive both immunotherapy and chemotherapy are excluded, because the cause of immune related adverse events cannot be properly identified.

### 2.6 Outcomes identification

The primary outcome of the study is the autoimmune-disease-free survival time after treatment of therapy, defined as the initiation date of immunotherapy or chemotherapy to the first record date of autoimmune-related diseases. Patients with no ICD records of irAEs are identified as having no events, and the at-risk (censoring) time is thereby defined as the treatment initiation time to the date of the last record of the lung cancer patient found in the database.

In the main analysis, the treatment initiation time is calculated regardless of the lines of the immunotherapy. A patient with irAEs is identified by consulting the corresponding ICD-9/ICD-10 codes of irAEs in the records. The types of autoimmune-related diseases are manually collected following expert suggestions and conventions of previous irAE studies, along with corresponding ICD-9 and ICD-10 codes from PheWAS, which are summarized in Supplementary Material Table S4. 56 categories of ICD codes relevant to irAEs are examined in this study, and the most common irAEs include Hypothyroidism, Rheumatoid arthritis, Myalgia (myositis) and thyroiditis. Due to the scope of this study, the definition of irAEs slightly discriminates from other studies. Some less severe autoimmune syndromes unlisted in the code list, such as skin rash and fatigue [25], are excluded from the study. The observational data is right censored at 3 years. Diagnosis after 3 years of initial treatment will be considered irrelavant to the therapy treatment.

In the main analysis any autoimmune diseases in the code list are considered as irAE. Section 4.3.2 provides sensitivity analysis results for four particular types of diseases: hypothyroidism, thyroditis, myalgia and myositis and rheumatoid arthritis. Other types of autoimmune diseases also exist in the records, but the sample size is not adequate for sensitivity analysis.

## 3 Methods

### 3.1 Cox proportional hazards model

Cox proportional hazards model is a popular technique used for survival-time (time-to-event) outcomes modeling on one or more predictors. Cox proportional hazards model does not require any assumptions about the distribution of event times, only assuming that:

1. Survival curves for different strata must have hazard functions that are proportional over the time *t*,
2. The relationship between the log hazard and each covariate is linear, which can be verified with residual plots.

The response variable *λ*(*t*) is defined as a hazard function which assesses the probability that the event of interest (in this study, this is defined as the time span from the first treatment to the first irAE diagnosis) occurred before *t*. The equation models this hazard as an exponential function of an arbitrary baseline hazard *λ*_0_ when all covariates are null, and *β* is the regression coefficient of all covariates *X*:

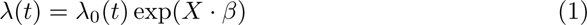

In the paper, standard log-rank test is also used to evaluate whether survival functions of the immunotherapy and chemotherapy groups are equal. The log-rank test is a statistical test used to compare the survival distributions of two or more groups. It is commonly employed in clinical trials and epidemiological studies to assess whether there are significant differences in the time to an event (such as death, disease progression, or failure) between groups.

### 3.2 Matching method

In this study, the treatment group is defined as patients who only receive immunotherapy, and the control group contains the patients who only receive chemotherapy. We apply matching method to assign higher weight to chemotherapy patients who have high similarity to each immunotherapy patients, following existing causal inference methods [26].

Matching methods efficiently alleviates the potential imbalance issue between the treatment and control group. In this study, each qualified patient that receives immunotherapy is matched to all ‘similar’ patients in the control group. A patient is considered ‘similar’ to another patient if all matching criteria are met:

1. The same sex, race and ethnicity, if the information is available without contradictory records. In the case of contradiction, i.e., when male and female coexist in the record of a patient, we mark the patient as ‘ambiguous’. An ambiguous patient will only be matched to ambiguous patient in the control group, following [26].
2. Age stratification. The difference in age of two matched patients should be no more than 2 years.
3. The same level of hospital visit rate. Percentile stratification in hospital visit rate is widely used in literature as an indicator of healthcare resource utilization pattern [27]. Hospital visit rate is defined as the average frequency of hospital visit (according to the record) during one year before the first diagnosis of lung cancer, which reveals the healthcare utilization of a patient. The healthcare utilization is classified into three categories: low (less than 67% quantile), medium (33% quantile to 67% quantile) and high (more than 33% quantile). Only patients with the same level of healthcare utilization can be matched.
4. Similar disease severity. In this study qualitative evaluation of disease severity is utilized as the matching criteria for previous disease burden. Two indicators are considered for defining similarity of disease burden: number of unique ICD codes found in the record prior to the first diagnosis of cancer, which quantifies the overall health condition of patients, as well as expert annotation. The level of disease severity is stratified into three categories: low (less than 67% quantile), medium (33% quantile to 67% quantile) and high (more than 33% quantile). Only patients with the same level of disease burden will be matched.
5. Exposure of treatment. The number of treatment cycles and the amount of exposure to the drug can be an important risk factor of irAEs. Therefore, the exposure of chemo- and immunotherapy for matched patients should be comparable. Guidance for dosage (exposure) comparison can be found in [24, 27].

Each patient in the control group may be selected multiple times as matched samples for different patients with immunotherapy. After acquiring the matched samples, we reweight the matched samples by inverse probability weighting [26]. Every patient in the treatment group is assigned with 1. If a treatment group patient has *n* matched samples, each sample will be assigned with an equal weight of ^1^.

After the re-weighting procedure the risks of irAEs among patients in two groups are compared, We conduct a survival analysis between two groups, along with the 95% confidence intervals. We first plot the Kaplan-Meier curve of two groups separately for original samples (before matching procedure) and matched samples, and test the difference of survival time distribution from irAEs. We further calculate the 95% confidence interval of hazard ratio. For pre-match samples, we apply Greenwood formula [28] for interval estimation. For matched samples, we apply Bootstrap method for interval estimation [29].

## 4 Results

### 4.1 Results overview

5725 lung cancer patients who received immune checkpoint inhibitors without prior autoimmune diseases are identified from cms database, with treatment initiation date ranging from 2011 to 2018. 2236 patients received Pembrolizumab, 132 received Atezolizumab, 3217 were on Novolumab and 230 were on Ipilimumab. For comparison froup, 102490 patients were identified who only received chemotherapy without prior autoimmune diagnosis. The distribution of immunotherapy and chemotherapy patients do not differentiate significantly in age or gender. 98.5% of the patients with immune checkpoint inhibitors can be matched to at least one sample from chemotherapy patients.

The distribution of immunotherapy treatments is presented in table 1. Pembrolizumab and Nivolumab has been increasingly popular after 2013, which has been confirmed in existing literature [30]. Although novel checkpoint inhibitors evolve each year, their application is limited, constituting a small percentage in the data.

### 4.2 Risk comparison of irAE

The Kaplan-Meier survival plot for immunotherapy in comparison to chemotherapy is shown in Figure 2. In the pre-matching analysis, we utilize all the samples who are qualified for this study, with vanilla log-rank test and cumulative incidence rate calculation. In post-matching analysis, adjusted samples are acquired through matching procedure and are assigned with corresponding weights (Section 3). Hazard ratio and accumulative incidence rate is shown in Table A11. According to both unadjusted (pre-matching) and adjusted (post-matching) samples, immunotherapy for lung cancer patients was associated with significantly higher risk of irAEs (*p* < 0.005).

**Fig. 2:**
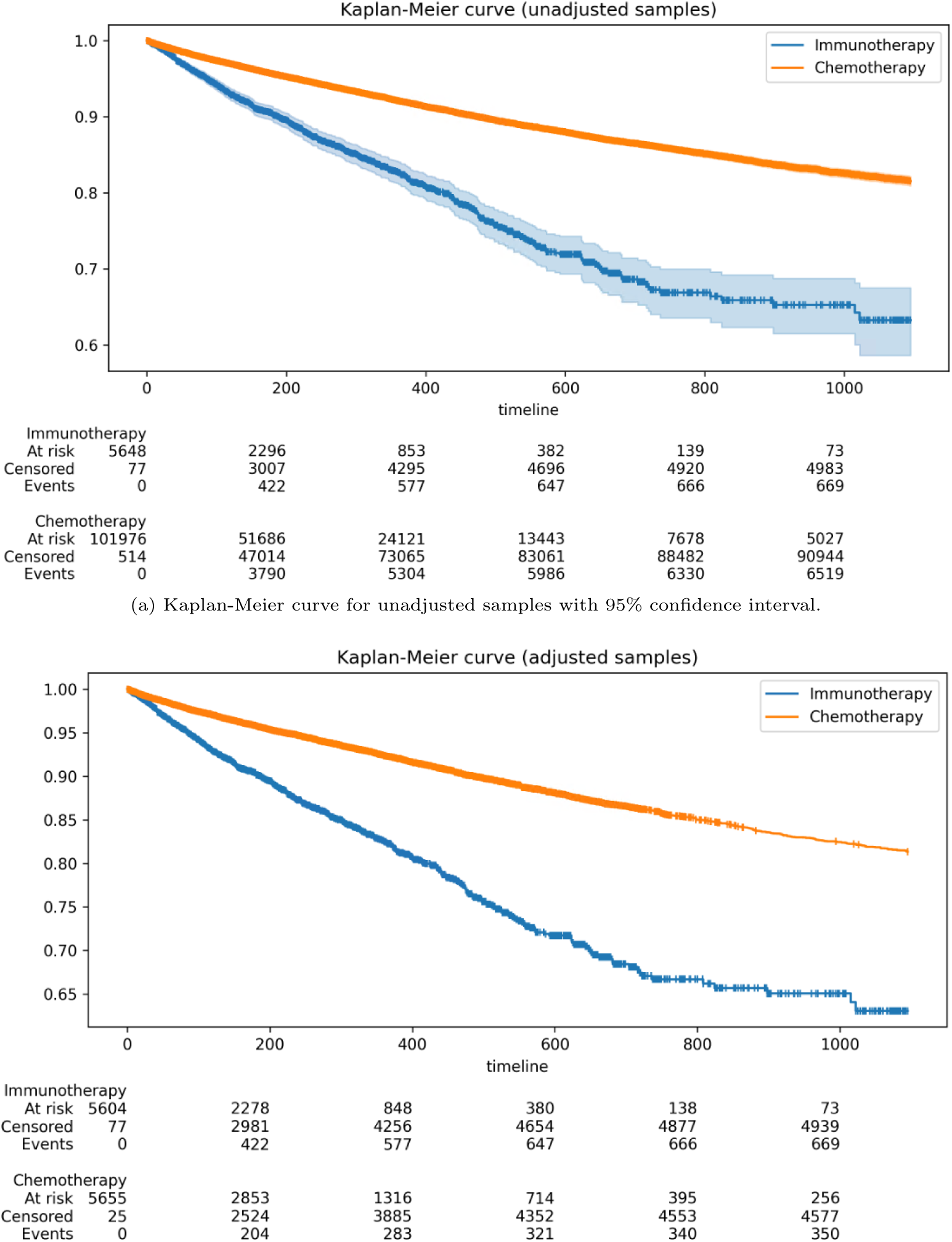
Kaplan-Meier curve for unadjusted and adjusted samples through time (in days).

### 4.3 Sensitivity analysis

We conducted several additional sensitivity analyses to illustrate the robustness of our conclusion.

To investigate the irAEs of different immunotherapy type, we conduct a drug-specific analysis on each type of immunotherapy, including Pembrolizumab, Nivolumab, Atezolizumab and Ipilimumab (see section 4.3.1). If a patient receives combination immunotherapy, they will be counted once in each subgroup of specific treatment they receive.

Immune checkpoint inhibitor may increase the risk of certain type of diseases [31]. To validate this we additionally investigated the increased risk of immune checkpoint inhibitors on four major types of irAEs: hypothyroidism, thyroiditis, rheumatoid arthritis and myalgia (see Section 4.3.2).

We also conducted stratified sensitivity analysis. In the main analysis, we set a 30-day quiescence, indicating the diagnosis of lung cancer must be at least 30 days after the first medical record date. We extensively investigate 60 days and 90 days as quiescence periods in the Supplementary Material Table S3.

#### 4.3.1 Therapy-specific analysis

In this section, we quantitively compare irAEs among different immunotherapies. Four main immunotherapy treatments are considered in this study: Ipilimumab Yervoy, Pembrolizumab Keytruda, Nivolumab Opdivo and Atezolizumab Tecentriq. While there are also other types of immunotherapy, the sample size is not enough for therapy-specific analysis. The Kaplan-Meier curves for unadjusted and adjusted samples are shown in Fig 3. Details of cumulative incidence ratio (CIR) are shown in Table 4.

**Fig. 3:**
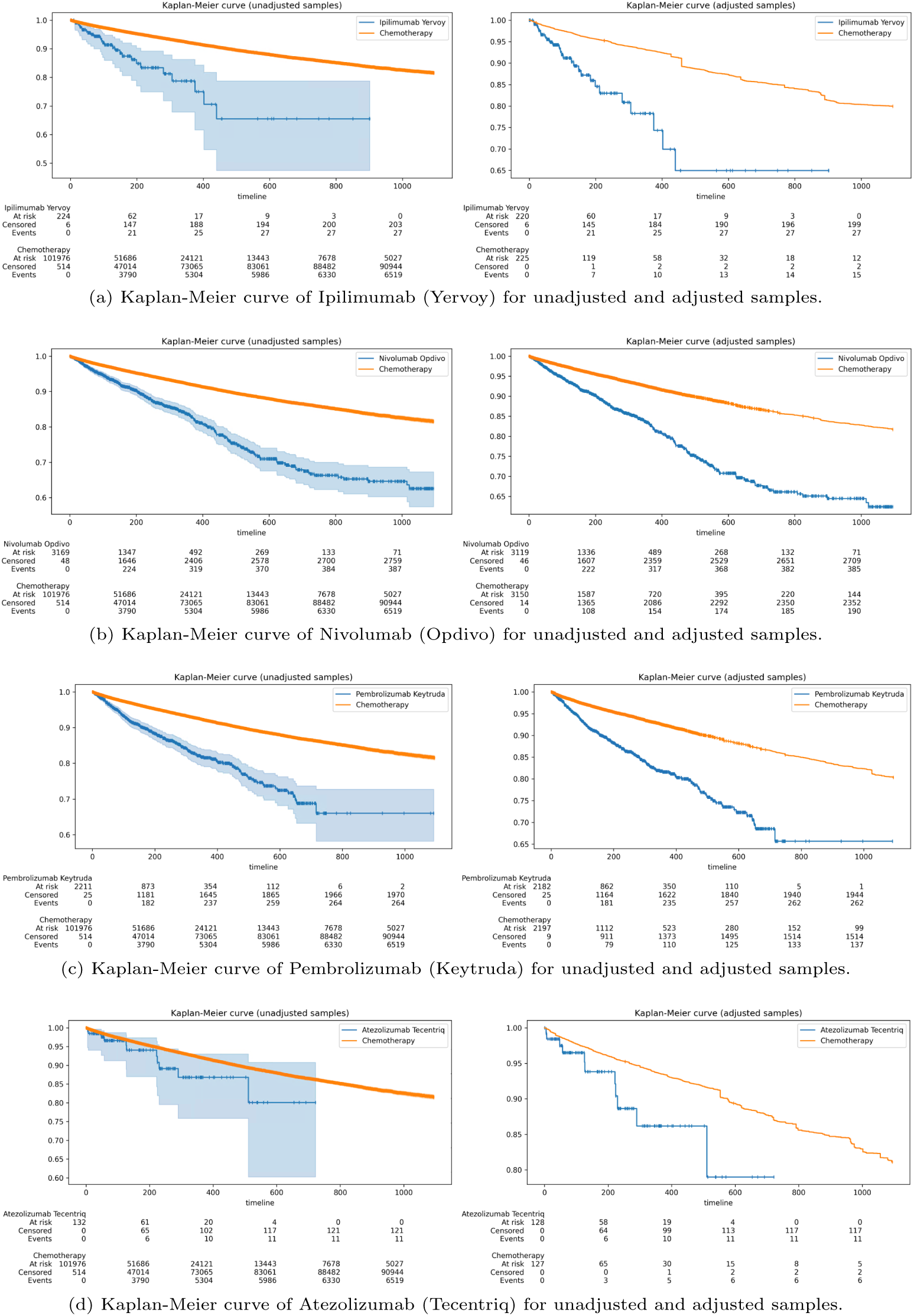
Therapy-specific analysis for unadjusted (left) and adjusted (right) samples.

**Fig. 4:**
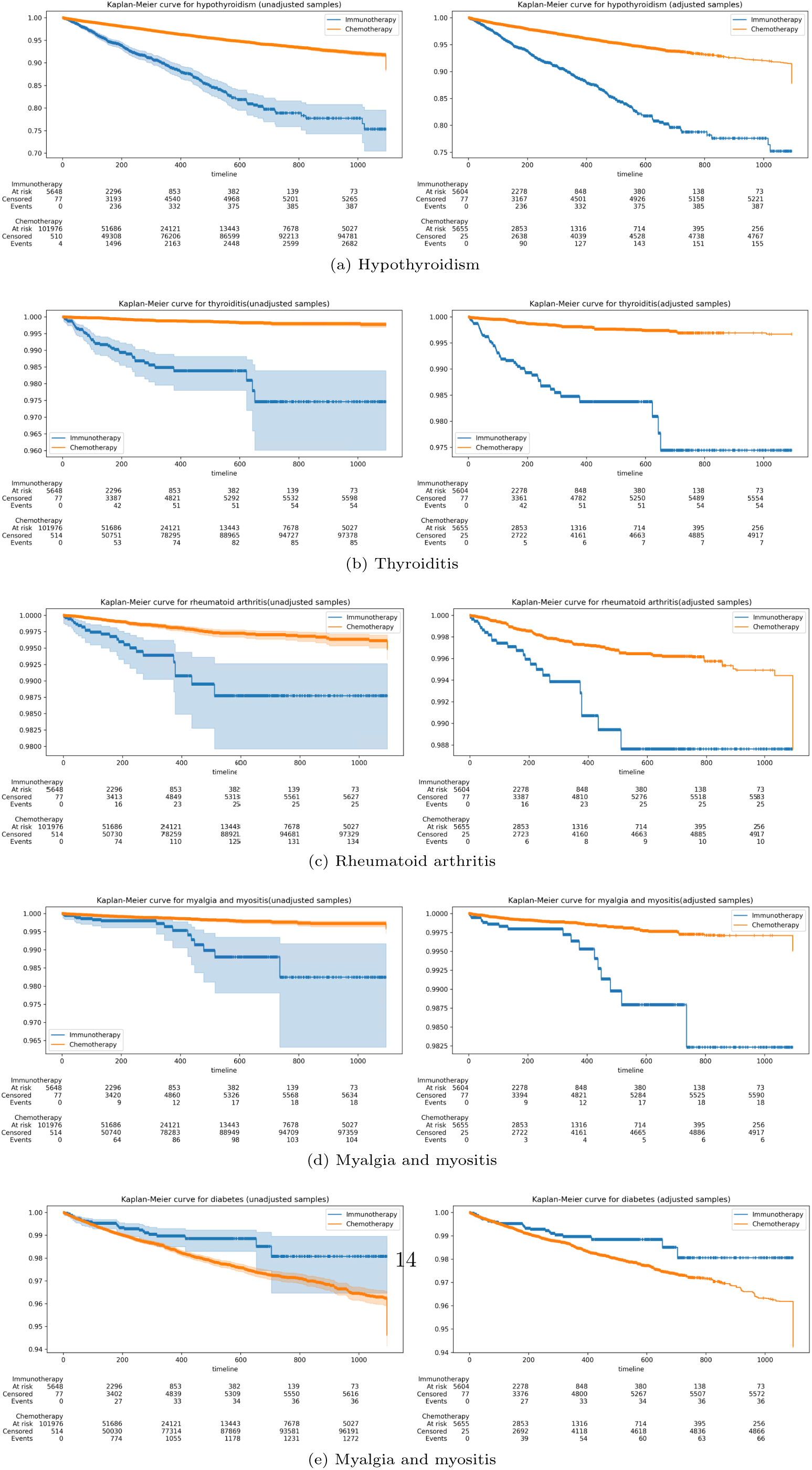
Disease-specific analysis for unadjusted (left) and adjusted (right) samples.

**Table 4:**
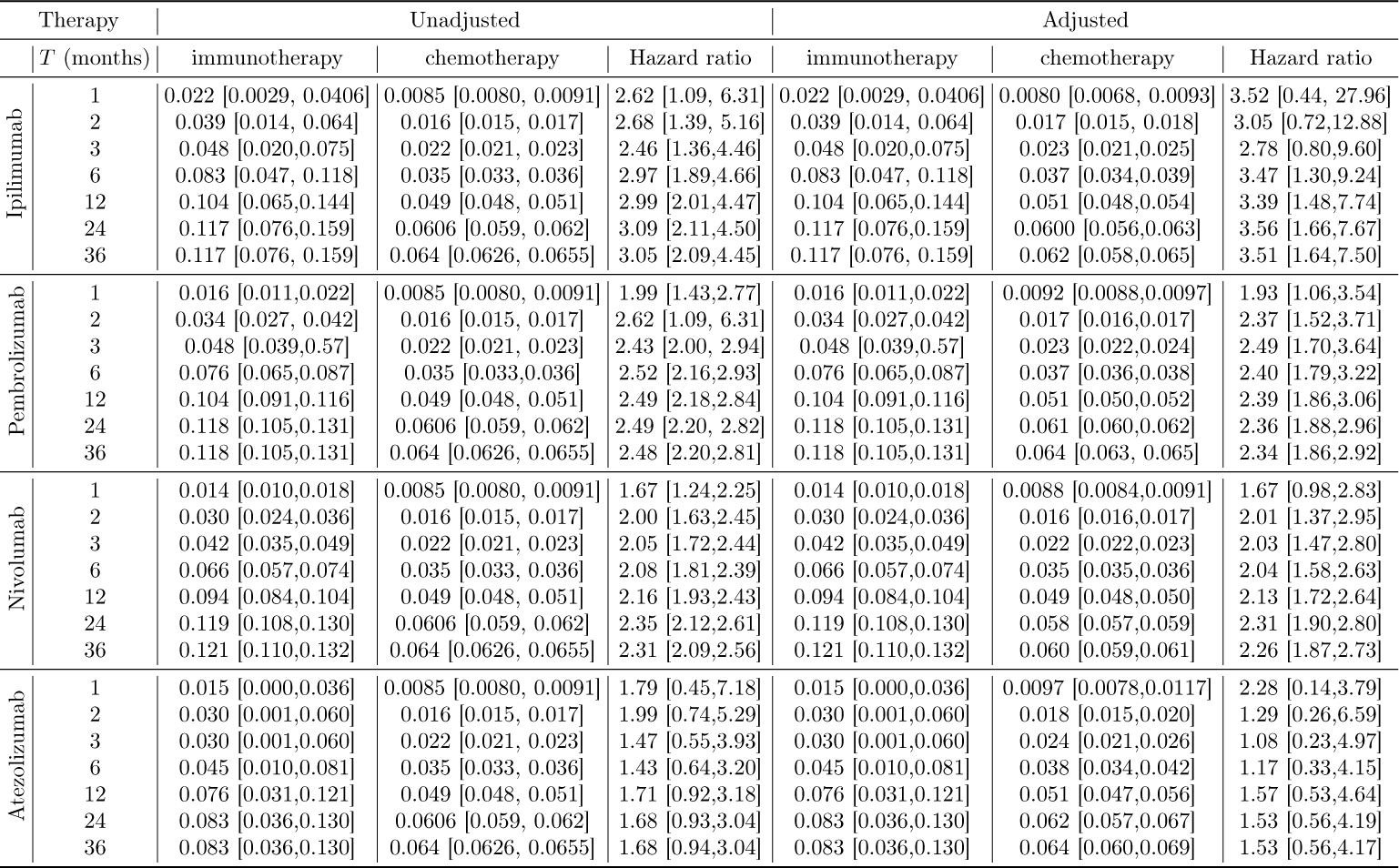
Cumulative incidence rate and hazard ratio.

#### 4.3.2 Disease-specific analysis

Existing literature has revealed that immunotherapy may increase the risk of certain autoimmune-related diseases. In this section we explore the irAEs of immunotherapy on five main categories of irAEs from the observations: diabetes [32], hypothyroidism [33], Hashimoto’s thyroiditis [34], rheumatoid arthritis [35] and myalgia and myositis [36]. While the adverse effect of diabetes is not statistically significant, the other four diseases are shown to be significantly related to immunotherapy. Table 5 shows the detailed results for unmatched and matched samples.

**Table 5:**
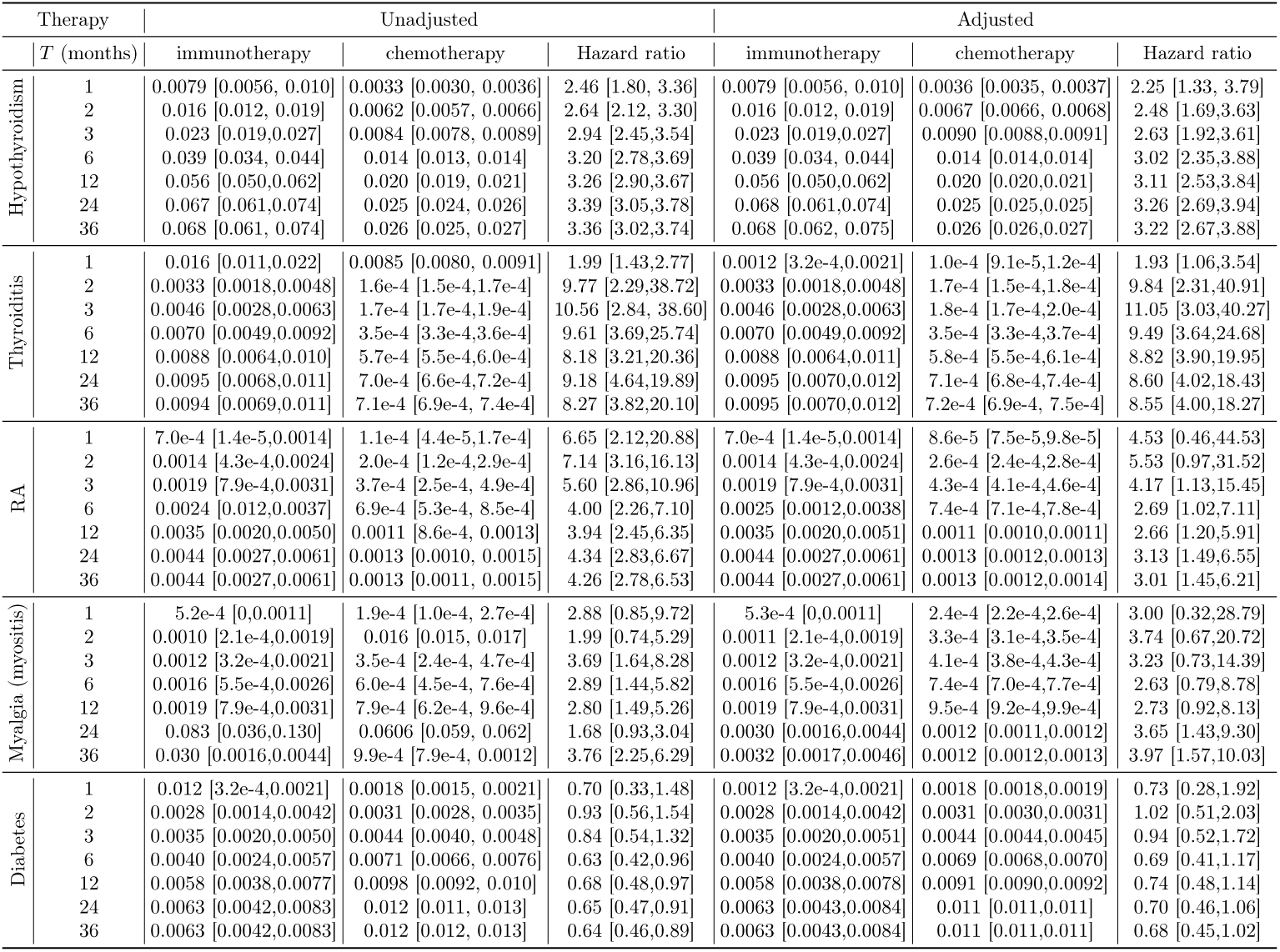
Cumulative incidence rate and hazard ratio.

### 4.4 Additional analysis

#### 4.4.1 Sub-population analysis

This study additionally analyzes irAEs among different subsets of study populations. We compared the risk of irAEs across different age groups. Comparison across different age groups in Supplementary Material Table S2 reveals that found that the irAE is statistically significant across all age groups, especially for patients of age 30 to 40.

#### 4.4.2 Sensitivity on quiescence time

In the main analysis, the quiescence time is fixed to 30 days, which indicates that any patient with no prior medical history at least 30 days before the first diagnosis of lung cancer will be excluded from the study. By further setting the time to 60 days, 90 days and 120 days, we explore whether the conclusion is robust to the selection of the period length of quiescence, with results shown in Table S3. The results illustrates the robustness of the conclusion to selection criteria of patients.

#### 4.4.3 Sensitivity on matching criteria

To balance the covariates of treatment group and control group, two variables are defined to quantify the overall health condition and the frequency of hospital visit: the total number of disease types of a patient one year prior the first diagnosis of lung cancer (sickness) and the number of hospital visits one year prior to the first diagnosis (hospital utilization). In the main study both variables are stratified into 3 levels by 33.3% and 66.6% quantile, and only patients with identical level of sickness and hospital utilization will be matched. Table S11 shows the results using different stratify strategies, which indicates that the results is insensitive to the matching criteria.

## 5 Discussion

In this study we conducted a large-scale cohort study to analyze the risk of autoimmune dieases associated with immuno checkpoint therapy on lung cancer patients. Results have shown that patients who received immunotherapy has patients with immunotherapy have a higher risk of developing irAEs, in comparison with lung cancer patients who receive chemotherapy. Specifically, certain immunotherapy treatments are associated with higher risk of autoimmune-related adverse effects, and risk of certain diseases is significantly higher than chemotherapy. Our result confirm the results of existing literature [37] and reveal the association of current immunotherapy drugs and the autoimmune-related adverse effects. Our result also suggests the slight difference in the presence of autoimmune syndromes among different immune checkpoint inhibitors, possibly due to difference of action mechanisms at PD-1 and PD-L1.

Thanks to the population size of lung cancer patients in the CMS source data, we conduct a significantly larger scale cohort study and explore autoimmune-related adverse effects from real world data compared to randomized controlled trials, which enables the discovery of certain diseases with very low incidence rate. Through a matching procedures, we investigate the causal relation between immunotherapy and the risk of autoimmune disorder with greater statistical power. Besides, our cohort study is broadly representative of real-world patients compared to the extrapolation from randomized controlled trial results, thus indicating more external validity [37]. An additional sensitivity analysis further shows that the risk of irAEs brought by immunotherapy is significant among different gender and age groups, which illustrates the robustness of our conclusion.

Our method complements the existing approaches of investigating irAEs of cancer therapies, and can be seen as a general methodology that can be extended to other treatment and disease phenotype researches. With specific drug (procedure) codes and corresponding ICD codes, our approach can evaluate any treatment type of interest for specific disease types, and has great potential for post-market adverse effect monitoring of FDA approved drugs. The sensitivity analyses conducted in the paper are also crucial for assuring the credibility of revealed adverse effects among different populations, i.e., age groups and genders.

There are also limitations in this study. First, to conduct the analysis, the diagnosis codes of irAEs must be pre-specified, while in the medical records, some less severe symptoms, such as fatigue and nausea, are not encoded to ICD codes properly, resulting in an potential inductive bias. Second, this study can only identify the date of diagnosis of certain disease, instead of the exact time of disease development. Therefore, some patients who have developed low-level autoimmune syndromes but were not clinically diagnosed, either due to inadequate syndromes or inaccessibility to hospital visits. Therefore, these patients cannot be accurately captured in the study. Besides, the ICD codes do not contain sufficient details that differentiate sub-categories of diseases or therapies, for example, histological sub-types and stage information of lung cancer or PD-L1 status. Another concern is the time difference of immunotherapy approval by FDA, and the approval of application on different cancer subtypes. For instance,

Nivolumab was approved by FDA for metastatic squamous non-small cell lung cancer in March 2015, and was later extended to metastatic non-squamous non-small cell lung cancer in October, 2015. In contrast, Atezolizumab was approved in October, 2016. Such time span may lead to evolution of chemotherapy treatments and time shifts of patient characteristics. However, such shifts are discarded in this study leading to potential inference bias. Lastly, the dataset cannot incorporate all lung cancer patients in the U.S. and the medicaid dataset mainly contains patients with relatively lower income, which may not accurately reflect the characteristics of the entire population. Further studies are still needed to verify our findings in other populations.

In conclusion, this study analyzes the risk of irAEs in lung cancer patients who receive immunotherapy, and suggests the necessity of clinically monitoring the autoimmune adverse effects in the patients with immune checkpoint inhibitors. Our results confirms the increased risk of autoimmune-related adverse effects due to immunotherapy compared to chemotherapy, and inspires further studies on the mechanism of such risk and monitoring of other adverse effects of immunotherapy for lung cancer patients.

## Conflict of Interest Statement

The authors declare that the research was conducted in the absence of any commercial or financial relationships that could be construed as a potential conflict of interest.

## Author Contributions

Yan Sun and Shihao Yang conceived and planned the protocol of the study. Yan Sun and Yunfan Guan carried out the analysis and contributed to the interpretation of the results. Yan Sun took the lead in writing the manuscript. All authors provided critical feedback and helped shape the research, analysis and manuscript.

## Funding

Research reported in this publication was supported by the National Institute of Diabetes and Digestive and Kidney Diseases of the National Institutes of Health under Award Number P30DK111024.The content is solely the responsibility of the authors and does not necessarily represent the official views of the National Institutes of Health.

## Data Availability Statement

The introduction of datasets analyzed for this study can be found at: https://www.ccwdata.org/web/guest/data-dictionaries.

## Appendix A Supplementary Tables and Figures

**Table A1:** HCPCS codes for immunotherapy.

| Chemotherapy name | Code | Code type | Description |
| --- | --- | --- | --- |
| CARBOPLATIN | J9045 | HCPC | Carboplatin, 50 mg |
| CISPLATIN | J9062 | HCPC | Cisplatin, 50 mg |
| CISPLATIN | C9418 | HCPC | CISPLATIN, POWDER OR SOLUTION, BRAND NAME, PER 10 MG |
| CISPLATIN | J9060 | HCPC | Cisplatin, powder or solution, per 10 mg |
| CISPLATIN | 209622 | HCPC | REIMB CISPLATIN 10 MG VIAL (PLATINOL). |
| DOCETAXEL | J9170 | HCPC | Docetaxel, 20 mg |
| DOCETAXEL | J9171 | HCPC | Injection, docetaxel, 1 mg |
| ETOPOSIDE | J9181 | HCPC | Etoposide, 10 mg |
| ETOPOSIDE | J9182 | HCPC | Etoposide, 100 mg |
| ETOPOSIDE | C9425 | HCPC | ETOPOSIDE, BRAND NAME, 10 MG |
| ETOPOSIDE | C9414 | HCPC | ETOPOSIDE, ORAL, BRAND NAME, 50 MG |
| ETOPOSIDE | J8560 | HCPC | Etoposide; oral, 50 mg |
| GEMCITABINE | J9201 | HCPC | Gemcitabine HCl, 200 mg |
| IRINOTECAN | 81350 | CPT4 | IRINOTECAN TOXICITY, UGT1A1*28, GENOTYPING |
| IRINOTECAN | J9206 | HCPC | Irinotecan, 20 mg |
| MECHLORETHAMINE | J9230 | HCPC | Mechlorethamine HCl, (nitrogen mustard), 10 mg |
| METHOTREXATE | 83520 | CPT4 | METHOTREXATE LEVEL |
| METHOTREXATE | J9250 | HCPC | Methotrexate sodium, 5 mg |
| METHOTREXATE | J9260 | HCPC | Methotrexate sodium, 50 mg |
| METHOTREXATE | J8610 | HCPC | Methotrexate: oral, 2.5 mg |
| PACLITAXEL | J9264 | HCPC | Injection, paclitaxel protein-bound particles, 1 mg |
| PACLITAXEL | J9267 | HCPC | INJECTION, PACLITAXEL, 1 MG |
| PACLITAXEL | J9265 | HCPC | Paclitaxel, 30 mg |
| PACLITAXEL | C9431 | HCPC | PACLITAXEL, BRAND NAME, 30 MG |
| PEMETREXED | J9305 | HCPC | Injection, pemetrexed, 10 mg |
| PEMETREXED | C9213 | HCPC | INJECTION, PEMETREXED, PER 10 MG |
| VINBLASTINE | J9360 | HCPC | Vinblastine sulfate, 1 mg |
| VINORELBINE | C9440 | HCPC | VINORELBINE TARTRATE, BRAND NAME, PER 10 MG |
| VINORELBINE | J9390 | HCPC | Vinorelbine tartrate, per 10 mg |

**Table A2:** 3-year cumulative incidence rate and hazard ratio for stratified age groups.

| Unmatched samples |  |  |  |
| --- | --- | --- | --- |
|  | CIR (immunotherapy) | CIR (chemotherapy) | Hazard ratio |
| ≤ 30 | 0.041 [0,0.087] | 0.044 [0.036,0.052] | 1.88 [0.59,5.97] |
| (30, 40] | 0.14 [0.054,0.22] | 0.059 [0.049,0.069] | 3.42 [1.74,6.73] |
| (40, 50] | 0.13 [0.10,0.17] | 0.067 [0.063,0.071] | 2.82 [2.16,3.68] |
| (50, 60] | 0.12 [0.10,0.13] | 0.067 [0.065,0.070] | 2.21 [1.93,2.54] |
| > 60 | 0.12 [0.10,0.13] | 0.061 [0.059,0.063] | 2.36 [2.12,2.63] |
| Matched samples |  |  |  |
|  | CIR (immunotherapy) | CIR (chemotherapy) | Hazard ratio |
| ≤ 30 | 0.039 [0,0.083] | 0.042 [0.034,0.050] | 1.83 [0.55,6.01] |
| (30, 40] | 0.13 [0.052,0.23] | 0.061 [0.049,0.069] | 3.40 [1.71,6.72] |
| (40, 50] | 0.13 [0.10,0.17] | 0.066 [0.062,0.073] | 2.84 [2.15,3.69] |
| (50, 60] | 0.12 [0.10,0.13] | 0.068 [0.066,0.072] | 2.20 [1.90,2.57] |
| > 60 | 0.13 [0.10,0.13] | 0.064 [0.058,0.065] | 2.34 [2.10,2.65] |

**Table A3:** 3-year cumulative incidence rate and hazard ratio for different quiescence time threshold.

| Unmatched samples |  |  |  |
| --- | --- | --- | --- |
| Quiescence time | CIR (immunotherapy) | CIR (chemotherapy) | Hazard ratio |
| 30 | 0.117 [0.109, 0.126] | 0.064 [0.063, 0.066] | 2.36 [2.18, 2.56] |
| 60 | 0.112 [0.102, 0.133] | 0.063 [0.062,0.066] | 2.34 [2.12, 2.55] |
| 90 | 0.114 [0.107, 0.128] | 0.063 [0.063, 0.067] | 2.41[2.31, 2.62] |
| 120 | 0.110 [0.099, 0.131] | 0.061 [0.059,0.065] | 2.29 [2.12, 2.75] |
| Matched samples |  |  |  |
| Quiescence time | CIR (immunotherapy) | CIR (chemotherapy) | Hazard ratio |
| 30 | 0.119 [0.110, 0.127] | 0.059 [0.059, 0.060] | 2.44 [2.14, 2.79] |
| 60 | 0.113 [0.101, 0.131] | 0.057 [0.057, 0.058] | 2.46 [2.12, 2.82] |
| 90 | 0.115 [0.105, 0.127] | 0.056 [0.055, 0.057] | 2.38 [2.10,2.84] |
| 120 | 0.110 [0.098,0.132] | 0.054 [0.052, 0.063] | 2.42 [2.13,2.86] |

**Table A4-1:**
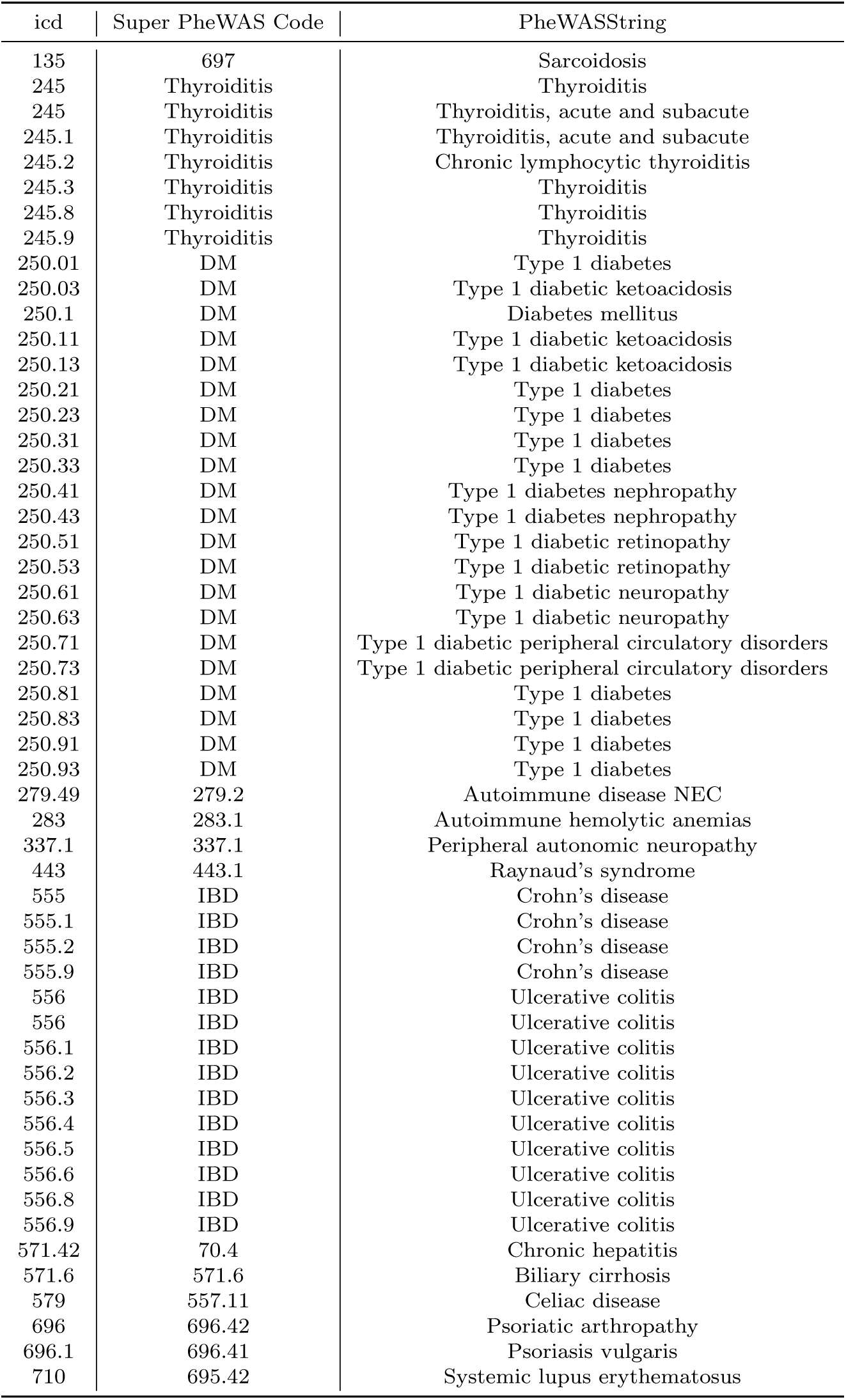
ICD-9 code for autoimmune diseases and syndromes included in the database (part 1).

| icd | Super PheWAS Code | PheWASString |
| --- | --- | --- |
| 135 | 697 | Sarcoidosis |
| 245 | Thyroiditis | Thyroiditis |
| 245 | Thyroiditis | Thyroiditis, acute and subacute |
| 245.1 | Thyroiditis | Thyroiditis, acute and subacute |
| 245.2 | Thyroiditis | Chronic lymphocytic thyroiditis |
| 245.3 | Thyroiditis | Thyroiditis |
| 245.8 | Thyroiditis | Thyroiditis |
| 245.9 | Thyroiditis | Thyroiditis |
| 250.01 | DM | Type 1 diabetes |
| 250.03 | DM | Type 1 diabetic ketoacidosis |
| 250.1 | DM | Diabetes mellitus |
| 250.11 | DM | Type 1 diabetic ketoacidosis |
| 250.13 | DM | Type 1 diabetic ketoacidosis |
| 250.21 | DM | Type 1 diabetes |
| 250.23 | DM | Type 1 diabetes |
| 250.31 | DM | Type 1 diabetes |
| 250.33 | DM | Type 1 diabetes |
| 250.41 | DM | Type 1 diabetes nephropathy |
| 250.43 | DM | Type 1 diabetes nephropathy |
| 250.51 | DM | Type 1 diabetic retinopathy |
| 250.53 | DM | Type 1 diabetic retinopathy |
| 250.61 | DM | Type 1 diabetic neuropathy |
| 250.63 | DM | Type 1 diabetic neuropathy |
| 250.71 | DM | Type 1 diabetic peripheral circulatory disorders |
| 250.73 | DM | Type 1 diabetic peripheral circulatory disorders |
| 250.81 | DM | Type 1 diabetes |
| 250.83 | DM | Type 1 diabetes |
| 250.91 | DM | Type 1 diabetes |
| 250.93 | DM | Type 1 diabetes |
| 279.49 | 279.2 | Autoimmune disease NEC |
| 283 | 283.1 | Autoimmune hemolytic anemias |
| 337.1 | 337.1 | Peripheral autonomic neuropathy |
| 443 | 443.1 | Raynaud's syndrome |
| 555 | IBD | Crohn's disease |
| 555.1 | IBD | Crohn's disease |
| 555.2 | IBD | Crohn's disease |
| 555.9 | IBD | Crohn's disease |
| 556 | IBD | Ulcerative colitis |
| 556 | IBD | Ulcerative colitis |
| 556.1 | IBD | Ulcerative colitis |
| 556.2 | IBD | Ulcerative colitis |
| 556.3 | IBD | Ulcerative colitis |
| 556.4 | IBD | Ulcerative colitis |
| 556.5 | IBD | Ulcerative colitis |
| 556.6 | IBD | Ulcerative colitis |
| 556.8 | IBD | Ulcerative colitis |
| 556.9 | IBD | Ulcerative colitis |
| 571.42 | 70.4 | Chronic hepatitis |
| 571.6 | 571.6 | Biliary cirrhosis |
| 579 | 557.11 | Celiac disease |
| 696 | 696.42 | Psoriatic arthropathy |
| 696.1 | 696.41 | Psoriasis vulgaris |
| 710 | 695.42 | Systemic lupus erythematosus |

**Table A4-2:**
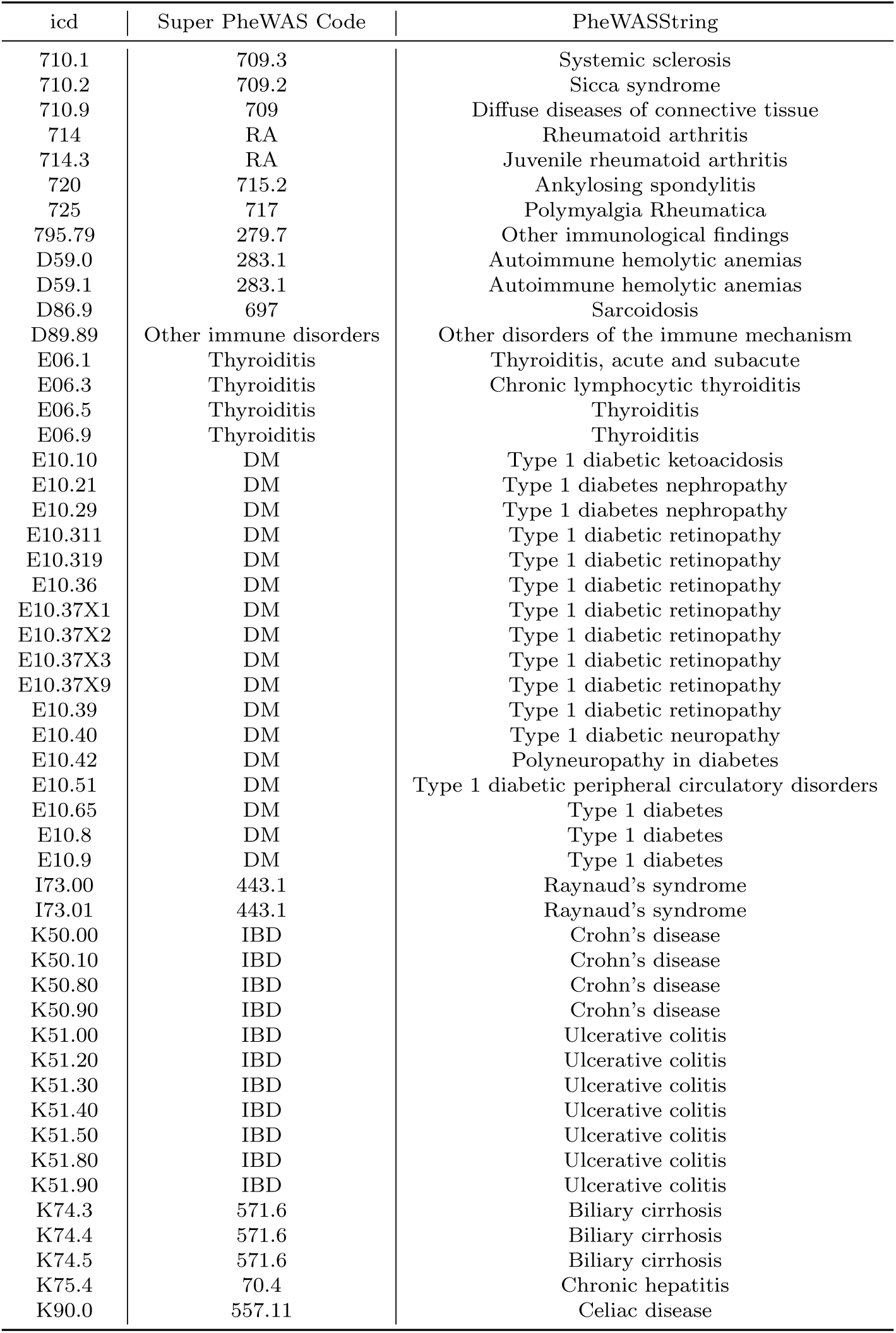
ICD-9 code for autoimmune diseases and syndromes included in the database (part 2).

**Table A4-3:** ICD-9 code for autoimmune diseases and syndromes included in the database (part 3).

| icd | Super PheWAS Code | PheWASString |
| --- | --- | --- |
| L40.0 | 696.41 | Psoriasis vulgaris |
| L40.1 | 696.41 | Psoriasis vulgaris |
| L40.2 | 696.41 | Psoriasis vulgaris |
| L40.3 | 696.41 | Psoriasis vulgaris |
| L40.4 | 696.41 | Psoriasis vulgaris |
| L40.50 | 696.42 | Psoriatic arthropathy |
| L40.54 | 696.42 | Psoriatic arthropathy |
| L40.59 | 696.42 | Psoriatic arthropathy |
| L40.8 | 696.41 | Psoriasis vulgaris |
| M06.9 | RA | Rheumatoid arthritis |
| M08.00 | RA | Juvenile rheumatoid arthritis |
| M32.10 | 695.42 | Systemic lupus erythematosus |
| M34.0 | 709.3 | Systemic sclerosis |
| M34.1 | 709.3 | Systemic sclerosis |
| M34.2 | 709.3 | Systemic sclerosis |
| M34.81 | 510.1 | Lung involvement in conditions classified elsewhere |
| M34.82 | 709.3 | Systemic sclerosis |
| M34.83 | 709.3 | Systemic sclerosis |
| M34.89 | 709.3 | Systemic sclerosis |
| M34.9 | 709.3 | Systemic sclerosis |
| M35.00 | 709.2 | Sicca syndrome |
| M35.01 | 709.2 | Sicca syndrome |
| M35.02 | 709.2 | Sicca syndrome |
| M35.03 | 709.2 | Sicca syndrome |
| M35.04 | 709.2 | Sicca syndrome |
| M35.09 | 709.2 | Sicca syndrome |
| M35.3 | 717 | Polymyalgia Rheumatica |
| M35.9 | 709 | Diffuse diseases of connective tissue |
| M45.9 | 715.2 | Ankylosing spondylitis |
| M60.9 | 770 | Myalgia and myositis NOS |
| M79.7 | 770 | Myalgia and myositis NOS |
| R76.8 | 279.7 | Other immunological findings |
| R76.9 | 279.7 | Other immunological findings |
| 99.3 | 711.2 | Reiter's disease |
| 136.1 | 711.3 | Behcet's syndrome |
| 242 | 242 | Thyrotoxicosis with or without goiter |
| 242.01 | 242.1 | Graves' disease |
| 242.9 | 242 | Thyrotoxicosis with or without goiter |
| 242.91 | 242 | Thyrotoxicosis with or without goiter |
| 244.9 | 244.4 | Hypothyroidism NOS |
| 255.41 | 255.21 | Glucocorticoid deficiency |
| 258.1 | 259 | Other endocrine disorders |
| 287.31 | 287.31 | Primary thrombocytopenia |
| 323.51 | Encephalitis | Encephalitis |
| 323.61 | Encephalitis | Encephalitis |
| 323.62 | Encephalitis | Encephalitis |
| 323.81 | Encephalitis | Encephalitis, non-infectious |
| 323.82 | Encephalitis | Encephalitis, non-infectious |
| 323.9 | Encephalitis | Encephalitis, non-infectious |
| 340 | 335 | Multiple sclerosis |
| 341 | Other CNS demyelinating diseases | Other demyelinating diseases of central nervous system |
| 341.8 | Other CNS demyelinating diseases | Other demyelinating diseases of central nervous system |
| 341.9 | Other CNS demyelinating diseases | Other demyelinating diseases of central nervous system |

**Table A4-4:**
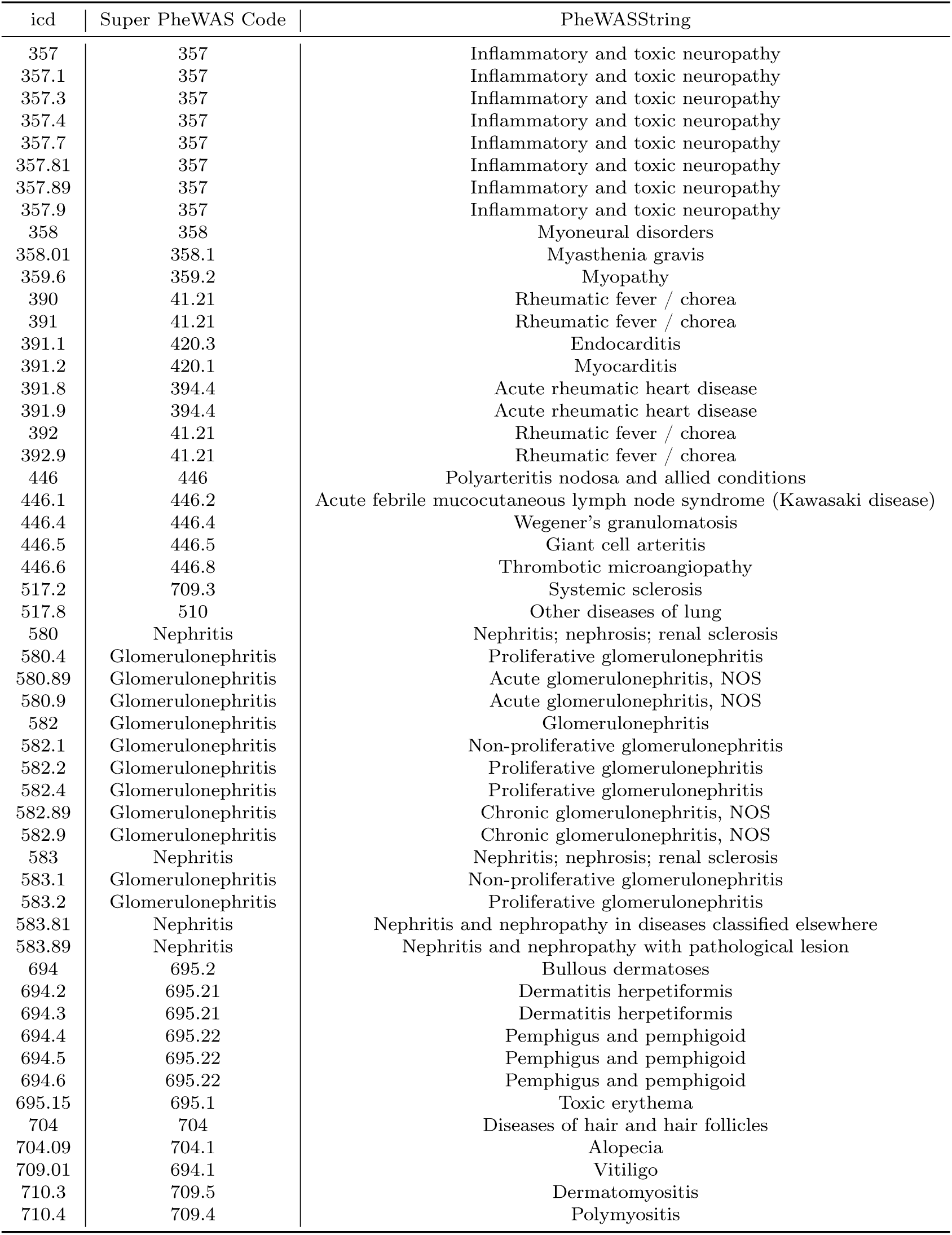
ICD-9 code for autoimmune diseases and syndromes included in the database (part 4).

| icd | Super PheWAS Code | PheWASString |
| --- | --- | --- |
| 357 | 357 | Inflammatory and toxic neuropathy |
| 357.1 | 357 | Inflammatory and toxic neuropathy |
| 357.3 | 357 | Inflammatory and toxic neuropathy |
| 357.4 | 357 | Inflammatory and toxic neuropathy |
| 357.7 | 357 | Inflammatory and toxic neuropathy |
| 357.81 | 357 | Inflammatory and toxic neuropathy |
| 357.89 | 357 | Inflammatory and toxic neuropathy |
| 357.9 | 357 | Inflammatory and toxic neuropathy |
| 358 | 358 | Myoneural disorders |
| 358.01 | 358.1 | Myasthenia gravis |
| 359.6 | 359.2 | Myopathy |
| 390 | 41.21 | Rheumatic fever / chorea |
| 391 | 41.21 | Rheumatic fever / chorea |
| 391.1 | 420.3 | Endocarditis |
| 391.2 | 420.1 | Myocarditis |
| 391.8 | 394.4 | Acute rheumatic heart disease |
| 391.9 | 394.4 | Acute rheumatic heart disease |
| 392 | 41.21 | Rheumatic fever / chorea |
| 392.9 | 41.21 | Rheumatic fever / chorea |
| 446 | 446 | Polyarteritis nodosa and allied conditions |
| 446.1 | 446.2 | Acute febrile mucocutaneous lymph node syndrome (Kawasaki disease) |
| 446.4 | 446.4 | Wegener's granulomatosis |
| 446.5 | 446.5 | Giant cell arteritis |
| 446.6 | 446.8 | Thrombotic microangiopathy |
| 517.2 | 709.3 | Systemic sclerosis |
| 517.8 | 510 | Other diseases of lung |
| 580 | Nephritis | Nephritis; nephrosis; renal sclerosis |
| 580.4 | Glomerulonephritis | Proliferative glomerulonephritis |
| 580.89 | Glomerulonephritis | Acute glomerulonephritis, NOS |
| 580.9 | Glomerulonephritis | Acute glomerulonephritis, NOS |
| 582 | Glomerulonephritis | Glomerulonephritis |
| 582.1 | Glomerulonephritis | Non-proliferative glomerulonephritis |
| 582.2 | Glomerulonephritis | Proliferative glomerulonephritis |
| 582.4 | Glomerulonephritis | Proliferative glomerulonephritis |
| 582.89 | Glomerulonephritis | Chronic glomerulonephritis, NOS |
| 582.9 | Glomerulonephritis | Chronic glomerulonephritis, NOS |
| 583 | Nephritis | Nephritis; nephrosis; renal sclerosis |
| 583.1 | Glomerulonephritis | Non-proliferative glomerulonephritis |
| 583.2 | Glomerulonephritis | Proliferative glomerulonephritis |
| 583.81 | Nephritis | Nephritis and nephropathy in diseases classified elsewhere |
| 583.89 | Nephritis | Nephritis and nephropathy with pathological lesion |
| 694 | 695.2 | Bullous dermatoses |
| 694.2 | 695.21 | Dermatitis herpetiformis |
| 694.3 | 695.21 | Dermatitis herpetiformis |
| 694.4 | 695.22 | Pemphigus and pemphigoid |
| 694.5 | 695.22 | Pemphigus and pemphigoid |
| 694.6 | 695.22 | Pemphigus and pemphigoid |
| 695.15 | 695.1 | Toxic erythema |
| 704 | 704 | Diseases of hair and hair follicles |
| 704.09 | 704.1 | Alopecia |
| 709.01 | 694.1 | Vitiligo |
| 710.3 | 709.5 | Dermatomyositis |
| 710.4 | 709.4 | Polymyositis |

**Table A4-5:** ICD-9 code for autoimmune diseases and syndromes included in the database (part 5).

| icd | Super PheWAS Code | PheWASString |
| --- | --- | --- |
| 710.8 | Other diffuse CNT diseases | Other specified diffuse diseases of connective tissue |
| 711.4 | Arthropathy | Arthropathy associated with infections |
| 711.41 | Arthropathy | Arthropathy associated with infections |
| 711.42 | Arthropathy | Arthropathy associated with infections |
| 711.43 | Arthropathy | Arthropathy associated with infections |
| 711.44 | Arthropathy | Arthropathy associated with infections |
| 711.45 | Arthropathy | Arthropathy associated with infections |
| 711.46 | Arthropathy | Arthropathy associated with infections |
| 711.47 | Arthropathy | Arthropathy associated with infections |
| 711.48 | Arthropathy | Arthropathy associated with infections |
| 711.49 | Arthropathy | Arthropathy associated with infections |
| 713.7 | Arthropathy | Arthropathy associated with other disorders classified elsewhere |
| 714.1 | RA | Rheumatoid arthritis |
| 714.2 | RA | Rheumatoid arthritis |
| 714.32 | RA | Juvenile rheumatoid arthritis |
| 714.81 | RA | Rheumatoid arthritis |
| 714.89 | RA | Rheumatoid arthritis and other inflammatory polyarthropathies |
| M02.30 | 711.2 | Reiter's disease |
| D86.0 | 697 | Sarcoidosis |
| M35.2 | 711.3 | Behcet's syndrome |
| E05.00 | 242 | Thyrotoxicosis with or without goiter |
| E05.01 | 242.1 | Graves' disease |
| E05.90 | 242 | Thyrotoxicosis with or without goiter |
| E05.91 | 242 | Thyrotoxicosis with or without goiter |
| E03.9 | 244.4 | Hypothyroidism NOS |
| E10.11 | DM | Type 1 diabetes |
| E10.618 | DM | Type 1 diabetes |
| E27.1 | 255.21 | Glucocorticoid deficiency |
| E31.0 | 259 | Other endocrine disorders |
| D69.3 | 287.31 | Primary thrombocytopenia |
| G04.02 | Encephalitis | Encephalitis |
| G04.00 | Encephalitis | Encephalitis |
| G04.30 | Encephalitis | Encephalitis |
| G04.81 | Encephalitis | Encephalitis, non-infectious |
| G04.89 | Encephalitis | Encephalitis, non-infectious |
| G04.90 | Encephalitis | Encephalitis, non-infectious |
| G35 | 335 | Multiple sclerosis |
| G36.0 | Other CNS demyelinating diseases | Other demyelinating diseases of central nervous system |
| G36.1 | Other CNS demyelinating diseases | Other demyelinating diseases of central nervous system |
| G36.9 | Other CNS demyelinating diseases | Other demyelinating diseases of central nervous system |
| G04.1 | 344 | Other paralytic syndromes |
| G61.0 | 357 | Inflammatory and toxic neuropathy |
| M05.50 | 357 | Inflammatory and toxic neuropathy |
| G13.1 | 357 | Inflammatory and toxic neuropathy |
| G61.1 | 357 | Inflammatory and toxic neuropathy |
| G61.81 | 357 | Inflammatory and toxic neuropathy |
| G61.89 | 357 | Inflammatory and toxic neuropathy |
| G61.9 | 357 | Inflammatory and toxic neuropathy |
| G70.00 | 358 | Myoneural disorders |
| G70.01 | 358.1 | Myasthenia gravis |
| M05.40 | 359.2 | Myopathy |
| I00 | 41.21 | Rheumatic fever / chorea |
| I01.0 | 41.21 | Rheumatic fever / chorea |

**Table A4-6:**
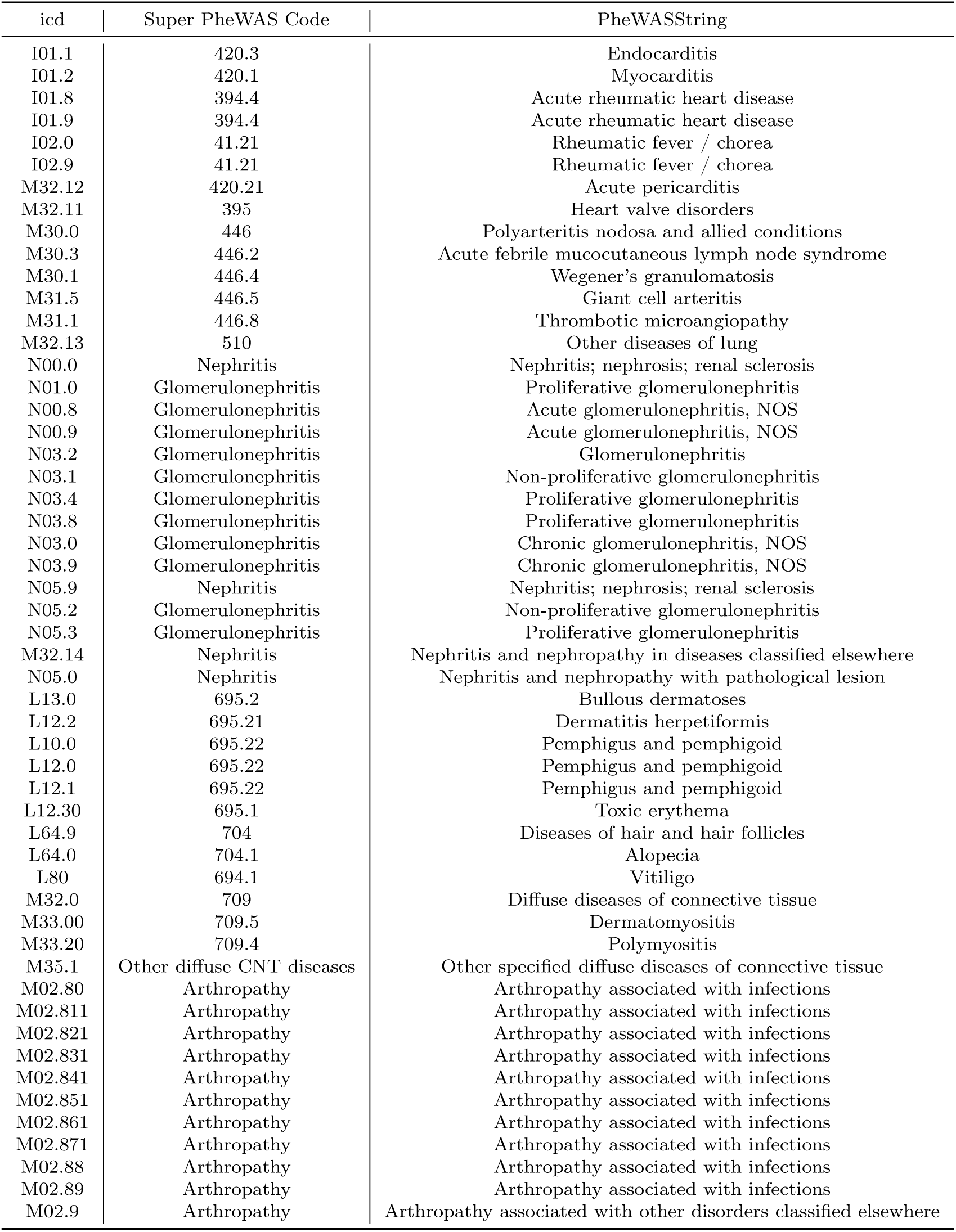
ICD-9 code for autoimmune diseases and syndromes included in the database (part 6).

**Table A4-7:** ICD-9 code for autoimmune diseases and syndromes included in the database (part 7).

| ICD | Super PheWAS Code | PheWASString |
| --- | --- | --- |
| M05.70 | RA | Rheumatoid arthritis and other inflammatory polyarthropathies |
| M05.00 | RA | Rheumatoid arthritis |
| M05.20 | RA | Rheumatoid arthritis |
| M08.40 | RA | Juvenile rheumatoid arthritis |
| M05.10 | RA | Rheumatoid arthritis |
| M06.4 | RA | Rheumatoid arthritis and other inflammatory polyarthropathies |
| M08.1 | 715 | Other inflammatory spondylopathies |
| E03.5 | 348.7 | Coma |

**Table A11:** Cumulative incidence rate and hazard ratio under different matching criteria.

| | $T$ (months) | CIR (immunotherapy) | CIR (chemotherapy) | Hazard ratio |
| --- | --- | --- | --- | --- |
| Bisection | 1 | 0.015 [0.012, 0.018] | 0.0083 [0.0081, 0.0086] | 1.77 [1.22, 2.53] |
|  | 2 | 0.032 [0.027, 0.036] | 0.016 [0.015, 0.016] | 2.18 [1.68, 2.86] |
|  | 3 | 0.045 [0.039, 0.050] | 0.020 [0.019, 0.022] | 2.31 [1.83, 2.89] |
|  | 6 | 0.070 [0.063, 0.076] | 0.034 [0.033, 0.035] | 2.32 [1.93, 2.77] |
|  | 12 | 0.098 [0.090, 0.106] | 0.046 [0.045, 0.047] | 2.41 [2.06, 2.76] |
|  | 24 | 0.118 [0.109, 0.126] | 0.054 [0.052, 0.057] | 2.51 [2.17, 2.87] |
|  | 36 | 0.119 [0.110, 0.127] | 0.061 [0.058, 0.062] | 2.47 [2.11, 2.82] |
| log-rank test | | $p < 0.01$ | | |
| Quadrisection | 1 | 0.015 [0.012, 0.018] | 0.0086 [0.0083, 0.0088] | 1.72 [1.20, 2.49] |
|  | 2 | 0.032 [0.027, 0.036] | 0.019 [0.017, 0.019] | 2.15 [1.63, 2.80] |
|  | 3 | 0.045 [0.039, 0.050] | 0.023 [0.022, 0.024] | 2.28 [1.83, 2.91] |
|  | 6 | 0.070 [0.063, 0.076] | 0.035 [0.035, 0.036] | 2.30 [1.91, 2.74] |
|  | 12 | 0.098 [0.090, 0.106] | 0.051 [0.049, 0.053] | 2.38 [2.03, 2.85] |
|  | 24 | 0.118 [0.109, 0.126] | 0.059 [0.057, 0.061] | 2.44 [2.13, 2.82] |
|  | 36 | 0.119 [0.110, 0.127] | 0.062 [0.059, 0.062] | 2.43 [2.11, 2.75] |
| log-rank test | | $p < 0.01$ | | |

## Footnotes

1 The number of patients in the category.

2 The proportion (in percentage) of patients in relation to the sub-population.

3 Some patients may receive combination therapy, which may be counted more than once in the subgroups.

4 Cumulative incidence rate

## Notes

### Competing Interest Statement

The authors have declared no competing interest.

### Funding Statement

This study was funded by NIH Award Number P30DK111024.

### Author Declarations

The study used ONLY openly available CCW data including medicare and medicaid research data that were originally located at:https://www.ccwdata.org/web/guest/data-dictionaries

